# Assessing hospital workers’ health: a tool for longitudinal health monitoring in a public tertiary hospital in Brazil’s Unified Health System (SUS)

**DOI:** 10.64898/2026.08.27.26361318

**Authors:** Rodrigo Teixeira Amancio, Letícia Nascimento Cruz, Rochelle da Silva Dantas, Márcia Pereira Gomes, Amanda de Araujo Batista da Silva, Pedro Emmanuel Alvarenga Americano do Brasil

## Abstract

**Background:** Health and administrative professionals in tertiary hospitals face high levels of occupational stress, mental illness, and multimorbidity. The integrated measurement of these multidimensional health aspects is essential for informing effective workplace health promotion strategies.

**Objective:** To describe the general health status of federal public hospital staff and correlate the measured health dimensions to inform institutional health promotion initiatives.

**Methods:** A cross-sectional, online survey study was conducted at Hospital Universitário dos Servidores do Estado (HUSE) between November and December 2025. Data collection was performed via online REDCap questionnaires covering sociodemographic profiles and validated instruments (SRQ-20, MIDAS, AUDIT, WHOQOL-BREF, PHI, WHOQOL-SRPB BREF, CBI, GPAQ, and EPSO). Descriptive statistics, comparisons across employment ties (permanent vs. contracted staff), and Spearman correlation matrices were calculated.

**Results:** Among 197 accesses, 117 completed the informed consent, and 86 finished all questionnaires. Participants were predominantly female, aged 40–60 years, and Christian. Screening positivity was 25% for common mental disorders, 21% for headache-related disability, and 10% for harmful drinking. Burnout scores clustered in the second quartile, while quality of life, happiness, and spirituality scores were in the upper third. Median physical activity was 670 min/week. Mental symptoms (SRQ-20), headache (MIDAS), and burnout (CBI) correlated positively with each other and negatively with quality of life, happiness, spirituality, and institutional support (EPSO).

**Conclusion:** The set of instruments proved feasible for situational health diagnosis among hospital staff. Although the sample size was limited in this baseline wave, the initiative fostered workplace health awareness, driving concrete initiatives, including an on-site functional gym and workplace vaccination campaigns.

**LAY SUMMARY:**

- ne in four hospital workers experienced significant mental health symptoms, and one in five suffered from headache-related disability, with burnout predominantly driven by work and personal stressors.
- levels of physical activity, spirituality, and perceived institutional support were correlated with lower burnout, reduced mental distress, and better overall quality of life.
- multi-domain digital questionnaires proved to be a practical tool for diagnosing workforce health, successfully inspiring immediate workplace health promotion actions such as planning an on-site gym and staff vaccination campaigns.

## INTRODUCTION

Health and administrative professionals working in general tertiary hospitals are exposed to a complex and demanding occupational environment that places a substantial burden on their physical, mental, and social well-being. Tertiary hospitals concentrate the most severe clinical cases, require continuous 24-hour operation, and demand high levels of technical expertise and emotional engagement from both clinical and non-clinical staff. Physicians, nurses, and allied health professionals face well-documented risks of burnout, depression, anxiety, and occupational stress, with studies identifying burnout prevalence exceeding 50% in public hospital networks and with administrative workers presenting higher odds of depression screening positivity than physicians.^1,2^ These conditions are not limited to clinical roles: service, administrative, and support staff also experience significant mental health burdens, compounded by financial strain, reduced job autonomy, and limited supervisory support.^1^ The compound effect of these stressors on the overall health of the hospital workforce represents a major public health and workforce sustainability challenge that demands systematic, evidence-based assessment.

Beyond mental health, hospital workers are vulnerable to a broad spectrum of chronic morbidities that frequently co-occur, constituting multimorbidity — defined as the simultaneous presence of two or more chronic conditions in a single individual. In the general working population, approximately half of workers report at least one chronic condition, and roughly one-quarter report multimorbidity, with conditions such as depression, musculoskeletal disorders, diabetes, cardiovascular disease, and hazardous alcohol use among the most prevalent.^3^ Longitudinal data show that approximately 5% of workers who initially had no chronic condition develop multimorbidity within five years, with risk heightened by non-standard employment, low job autonomy, and female sex.^4^ For hospital workers specifically, occupational exposures — including shift work, irregular schedules, physical demands, and psychosocial stressors — are known to accelerate the accumulation of chronic conditions, with multimorbidity in turn amplifying absenteeism, presenteeism, disability, and healthcare costs.^3^ Understanding the specific burden and distribution of multimorbidity in this population is therefore essential to designing effective, targeted health promotion strategies.

Adequate characterization of the health status of hospital workers requires the use of validated, multidimensional instruments capable of capturing the breadth of physical, mental, occupational, and spiritual health domains. The Self-Reporting Questionnaire-20 (SRQ-20) is a widely used WHO-endorsed screening tool for common mental disorders, validated in Brazilian populations. The Alcohol Use Disorders Identification Test (AUDIT) assesses hazardous and harmful alcohol use. The Migraine Disability Assessment (MIDAS) quantifies headache-related disability. The Copenhagen Burnout Inventory (CBI) measures personal, work-related, and client-related burnout across occupational groups, including non-clinical staff, and has been applied alongside the WHO Quality of Life instrument (WHOQOL-BREF) to characterize burnout and quality of life in hospital settings.^5^ The WHOQOL-SRPB BREF extends quality-of-life assessment to spirituality, religiosity, and personal beliefs — dimensions that may be particularly relevant in Brazilian cultural contexts. The Global Physical Activity Questionnaire (GPAQ), developed by WHO, captures physical activity across occupational, transport, and leisure domains. Together, these instruments offer a comprehensive, multi-domain picture of health status that no single instrument can provide alone, and their combined use reflects current recommendations for integrated, sector-specific assessment frameworks.^6^

The World Health Organization, through its Global Plan of Action on Workers’ Health (2008–2017) and subsequent frameworks, has consistently advocated for the workplace as a priority setting for health promotion, with hospitals explicitly recognized as settings in which healthy workplace programs should be implemented.^7^ Systematic reviews of hospital-based interventions confirm that multi-component strategies combining physical activity promotion, dietary improvement, financial incentives, and motivational approaches are the most effective in improving health behaviors among hospital staff, with 15 of 18 high- or moderate-quality studies reporting at least one positive health outcome.^7^ Whole-system approaches — encompassing all staff categories, visible leadership engagement, and adaptation to local workforce needs — have demonstrated statistically significant improvements in both subjective mental health and objectively measured physical health outcomes.^8^ Despite this evidence base, implementation of structured health promotion programs in Brazilian public hospitals remains limited, and comprehensive baseline data on the multidimensional health status of these workers are scarce, hampering the development of evidence-based, context-appropriate interventions.

Identifying and assessing multimorbidity—combined with an analysis of behavioral aspects such as burnout, spirituality, quality of life (QoL), and perceptions of institutional care—are fundamental steps toward informing prevention strategies, promoting well-being, and improving human resource management within the healthcare system. This research aims to describe the general health status of federal public hospital staff and to correlate the measured dimensions to discuss possible health promotion actions.

## METHOD

### Ethical aspects

The project was approved by the Research Ethics Committee of Hospital Universitário dos Servidores do Estado (HUSE) (former Hospital Federal dos Servidores do Estado - HFSE) (CAAE: 90857525.7.0000.5252; Approval No. 7.842.624, 17/09/2025), following CNS Resolution No. 466/2012.

### Data source and participants

The HUSE staff comprises two formal employment categories: permanent and contracted staff. Hospital staff were invited to participate in this research; thus, participation is voluntary, and no incentives were offered. HUSE workers were invited to participate via email and WhatsApp, based on the employment roster provided by the Human Resources department. Internal promotion—providing clarifications and information—was conducted using project-specific posters, duly authorized by the HFSE administration, before the start of data collection, including the project’s participation in the hospital’s 78th-anniversary celebration and a group walk for staff members, featuring the distribution of T-shirts and pins to promote the project.

When clicking on the invitation link, the respondents accessed a RedCap survey, where the first page was a brief project explanation with instructions, including the time length for filling out the forms (approximately 150 questions). Pressing “next page”, the respondents accessed a written consent form to sign, which was immediately sent to the informed e-mail and allowed the filling out of all the following research forms. Data collection was performed from November 11th, 2025, through December 12th, 2025. Respondents were not able to review their answers after a saved form, and they had the chance to start answering and finish at any time up to the end of the survey with a “returning code” provided by RedCap individually. Every entry was guaranteed to be unique for each respondent by e-mail and phone number.

The inclusion criteria were: (a) age 18 years or older; (b) HUSE staff with an active formal employment relationship. The exclusion criteria were: (a) inability to carry out any part of the protocol, in the researcher’s judgment, during the initial assessment, including functional illiteracy or cognitive impairment; (b) workers on medical leave, vacation, or unpaid leave during the data collection period.

### Evaluating procedures

There are various characteristics related to the individual, as well as their family and professional situation, including income, that were of interest, and several validated instruments were applied. The descriptive characteristics were mostly inspired by forms used by the *Instituto Nacional de Geografia e Estatística* (IBGE) in the Brazilian national census,^9,10^ and the validated applied questionnaires were the “Self Reporting Questionnaire” (SQR-20)^11^ to check mental symptoms, the “Migraine Disability Assessment” (MIDAS)^12^ to check disabilities due to headaches, the “Alcohol Use Disorders Identification Test” (AUDIT)^13,14^ to check alcohol consumption, the Brief WHO Quality of Life (WHOQOL-BREF)^15^ to check quality of life, the “Pemberton Happiness Index” (PHI)^16^ and Cantril ladder^17^ to check happiness, the “World Health Organization’s Quality of Life Instrument – Spirituality, Religion, and Personal Beliefs”

(WHOQOL-SRPB BREF)^18^ to check spirituality, the “Copenhagen Burnout Inventory” (CBI)^19^ to check burnout, the “Global Physical Activity Questionnaire” (GPAQ)^20^ to check physical activity, and the “Escala de Percepção de Suporte Organizacional” (EPSO)^21^ to check the institutional support to its employees.

### Analysis plan

The analysis plan consisted of quantitative descriptions (counts, frequencies, measures of central tendency, and measures of dispersion) of baseline information, such as sociodemographic characteristics. These characteristics may be compared across groups of interest using the Wilcoxon rank-sum test for continuous and ordinal variables and Fisher’s exact test or Pearson’s chi-squared test for categorical variables, selected according to the data’s format and expected cell frequencies. For the questionnaires measuring the aforementioned dimensions, total scores for each instrument were calculated and compared across the same groups of interest used to stratify the sociodemographic data, using the same hypothesis tests. Additionally, a Spearman correlation matrix was built among the total scores of the validated instruments to address the study’s aim of correlating the measured health dimensions. A two-sided p-value < 0.05 was considered statistically significant, with no correction applied for multiple comparisons. Analyses will be conducted using the R-project software.

## RESULTS

There were 197 accesses to the online survey, and 117 unique informed consents were completed. One hundred respondents completed the first survey form (socio-demographic characteristics), and 86 completed all forms. The third quantile time of filling each form was always less than 9 minutes, showing that this form combination is reasonably feasible. (Figure 1)

**Figure 1.**
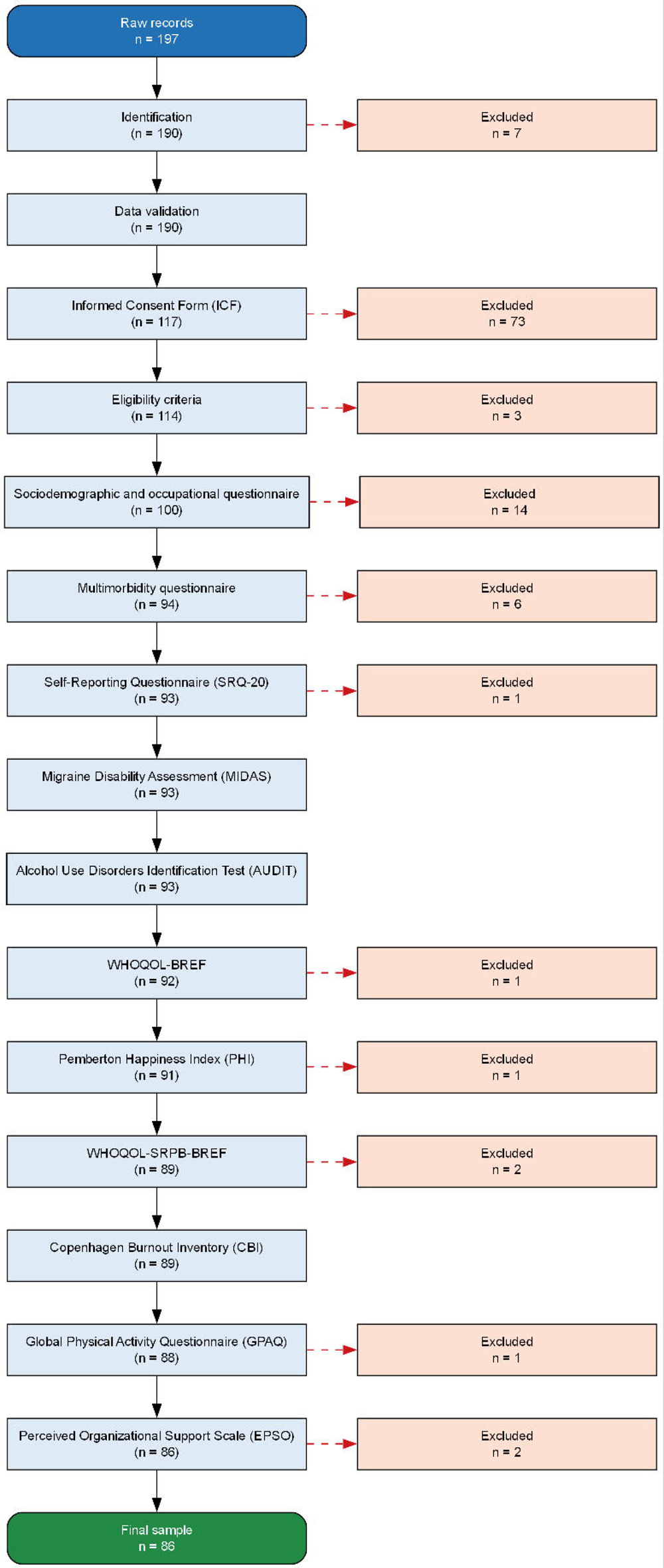
Flow of participants from raw survey records to the final analytic sample. At each stage, participants were retained only if the corresponding instrument was marked complete; those who did not complete it were excluded from the following stage (dashed arrows). Abbreviations: ICF, Informed Consent Form; SRQ-20, Self-Reporting Questionnaire; MIDAS, Migraine Disability Assessment; AUDIT, Alcohol Use Disorders Identification Test; WHOQOL-BREF, World Health Organization Quality of Life Instrument, abbreviated version; PHI, Pemberton Happiness Index; WHOQOL-SRPB-BREF, WHOQOL Spirituality, Religiousness, and Personal Beliefs module, abbreviated version; CBI, Copenhagen Burnout Inventory; GPAQ, Global Physical Activity Questionnaire; EPSO, Perceived Organizational Support Scale.

The respondents were mainly between 40 and 60 years old, female, permanent hospital staff, white-skinned people, and declared themselves Christians. The respondents’ income was widely different between the permanent and contracted staff, with a similar number of dependent households. The contracted staff were mainly administrative and cleaning personnel, while the permanent staff were mainly in the health field. The median time since graduation was higher, and the weekly working hours were lower among the permanent staff. The contracted staff were most dependent on public transportation, and the majority of respondents usually spent more than one hour commuting to work. (Table 1)

**Table 1.** Social, demographic, and morbidity sample characteristics by the hospital staff employment relationship.

| Variable | Employment status |  |  | p-value<sup>2</sup> |
| --- | --- | --- | --- | --- |
|  | Overall<br>N = 86<sup>1</sup> | Permanent staff<br>N = 57<sup>1</sup> | Contract staff<br>N = 21<sup>1</sup> |  |
| **Age (years)** | 47 (43, 53) | 48 (45, 54) | 43 (34, 52) | 0.005 |
| **Age group** |  |  |  | 0.004 |
| 18-29 | 5 (5.8%) | 1 (1.8%) | 4 (14%) |  |
| 30-39 | 7 (8.1%) | 1 (1.8%) | 6 (21%) |  |
| 40-49 | 39 (45%) | 29 (51%) | 10 (34%) |  |
| 50-59 | 26 (30%) | 20 (35%) | 6 (21%) |  |
| 60+ | 9 (10%) | 6 (11%) | 3 (10%) |  |
| **Sex at birth** |  |  |  | 0.6 |
| Female | 62 (72%) | 42 (74%) | 20 (69%) |  |
| Male | 24 (28%) | 15 (26%) | 9 (31%) |  |
| **Race/Skin color** |  |  |  | 0.2 |
| White | 48 (56%) | 33 (59%) | 15 (52%) |  |
| Brown | 25 (29%) | 13 (23%) | 12 (41%) |  |
| Black | 12 (14%) | 10 (18%) | 2 (6.9%) |  |
| Missing information | 1 | 1 | 0 |  |
| **Religion** |  |  |  | 0.2 |
| Catholic | 31 (36%) | 22 (39%) | 9 (31%) |  |
| Evangelical | 13 (15%) | 9 (16%) | 4 (14%) |  |
| Spiritist | 18 (21%) | 14 (25%) | 4 (14%) |  |
| No religion | 10 (12%) | 6 (11%) | 4 (14%) |  |
| Umbanda/Candomblé/Afro-Brazilian religions | 7 (8.2%) | 2 (3.6%) | 5 (17%) |  |
| Multiple religious affiliations | 2 (2.4%) | 1 (1.8%) | 1 (3.4%) |  |
| Jehovah's Witnesses | 1 (1.2%) | 0 (0%) | 1 (3.4%) |  |
| World Messianic Church | 1 (1.2%) | 1 (1.8%) | 0 (0%) |  |
| Does not know | 1 (1.2%) | 0 (0%) | 1 (3.4%) |  |
| Other Christian religious affiliations/Others | 1 (1.2%) | 1 (1.8%) | 0 (0%) |  |
| Missing information | 1 | 1 | 0 |  |
| **Education level** |  |  |  | <0.001 |
| High school or lower | 5 (5.9%) | 2 (3.5%) | 3 (11%) |  |
| Completed higher education | 39 (46%) | 18 (32%) | 21 (75%) |  |
| Graduate degree | 41 (48%) | 37 (65%) | 4 (14%) |  |
| Missing information | 1 | 0 | 1 |  |
| **Individual income (R\$)** | | | | <0.001 |
| 1.00 to 500.00 | 23 (27%) | 15 (27%) | 8 (28%) |  |
| 1,001.00 to 2,000.00 | 6 (7.1%) | 0 (0%) | 6 (21%) |  |
| 2,001.00 to 3,000.00 | 11 (13%) | 2 (3.6%) | 9 (31%) |  |
| 3,001.00 to 5,000.00 | 11 (13%) | 8 (15%) | 3 (10%) |  |
| 5,001.00 to 10,000.00 | 16 (19%) | 13 (24%) | 3 (10%) |  |
| 10,001.00 to 20,000.00 | 11 (13%) | 11 (20%) | 0 (0%) |  |
| 20,001.00 to 100,000.00 | 6 (7.1%) | 6 (11%) | 0 (0%) |  |
| Missing information | 2 | 2 | 0 |  |
| **Income in minimum wages** | 2.6 (0.0, 5.3) | 4.0 (0.0, 7.9) | 1.4 (0.0, 2.0) | <0.001 |
| Missing information | 2 | 2 | 0 |  |
| **Household income (R\$)** | 7,000 (2,750, 15,000) | 12,000 (5,000, 24,000) | 4,000 (2,000, 6,000) | <0.001 |
| Missing information | 1 | 1 | 0 |  |
| **Per capita household income (R\$)** | 3,000 (1,000, 7,000) | 5,000 (1,500, 10,000) | 1,667 (800, 2,695) | <0.001 |
| Missing information | 1 | 1 | 0 |  |
| **Dependent people in the household** |  |  |  | 0.3 |
| 1 | 15 (17%) | 9 (16%) | 6 (21%) |  |
| 2 | 26 (30%) | 17 (30%) | 9 (31%) |  |
| 3 | 26 (30%) | 18 (32%) | 8 (28%) |  |
| 4 | 15 (17%) | 12 (21%) | 3 (10%) |  |
| 5 | 4 (4.7%) | 1 (1.8%) | 3 (10%) |  |
| **Donated to charity** |  |  |  | 0.3 |
| Yes | 28 (33%) | 21 (37%) | 7 (25%) |  |
| No | 57 (67%) | 36 (63%) | 21 (75%) |  |
| Missing information | 1 | 0 | 1 |  |
| **Financial support network** |  |  |  | 0.7 |
| Yes | 57 (66%) | 37 (65%) | 20 (69%) |  |
| No | 29 (34%) | 20 (35%) | 9 (31%) |  |
| **Field of work** |  |  |  | <0.001 |
|  | Overall<br>N = 86<sup>1</sup> | Permanent staff<br>N = 57<sup>1</sup> | Contract staff<br>N = 21<sup>1</sup> |  |
| Administrative technician | 16 (20%) | 3 (5.8%) | 13 (46%) |  |
| Apprentice | 1 (1.3%) | 0 (0%) | 1 (3.6%) |  |
| Biologist | 4 (5.0%) | 1 (1.9%) | 3 (11%) |  |
| Cleaning assistant | 2 (2.5%) | 0 (0%) | 2 (7.1%) |  |
| Clinical laboratory technician | 2 (2.5%) | 2 (3.8%) | 0 (0%) |  |
| Maintenance officer | 1 (1.3%) | 0 (0%) | 1 (3.6%) |  |
| Nurse | 16 (20%) | 16 (31%) | 0 (0%) |  |
| Nursing technician | 10 (13%) | 10 (19%) | 0 (0%) |  |
| Nutrition and dietetics technician | 1 (1.3%) | 1 (1.9%) | 0 (0%) |  |
| Nutritionist | 5 (6.3%) | 3 (5.8%) | 2 (7.1%) |  |
| Occupational therapist | 2 (2.5%) | 2 (3.8%) | 0 (0%) |  |
| Other health-related areas | 1 (1.3%) | 1 (1.9%) | 0 (0%) |  |
| Other, non-health-related areas | 4 (5.0%) | 2 (3.8%) | 2 (7.1%) |  |
| Pharmacist | 2 (2.5%) | 2 (3.8%) | 0 (0%) |  |
| Physical therapist | 2 (2.5%) | 2 (3.8%) | 0 (0%) |  |
| Physician | 4 (5.0%) | 4 (7.7%) | 0 (0%) |  |
| Radiology technician | 3 (3.8%) | 3 (5.8%) | 0 (0%) |  |
| Receptionist | 1 (1.3%) | 0 (0%) | 1 (3.6%) |  |
| Secretarial technician | 3 (3.8%) | 0 (0%) | 3 (11%) |  |
| Missing | 6 | 5 | 1 |  |
| **Years since graduation** | 22 (14, 27) | 24 (21, 28) | 6 (2, 19) | <0.001 |
| Missing information | 6 | 1 | 5 |  |
| **Working hours per week** | 42 (30, 44) | 40 (30, 44) | 44 (40, 44) | 0.062 |
| Missing | 1 | 0 | 1 |  |
| **Means of transportation to work** |  |  |  | <0.001 |
| On foot | 1 (1.2%) | 1 (1.8%) | 0 (0%) |  |
| Bus | 31 (36%) | 15 (26%) | 16 (55%) |  |
| BRT (Bus Rapid Transit) | 4 (4.7%) | 1 (1.8%) | 3 (10%) |  |
| Train or subway | 24 (28%) | 15 (26%) | 9 (31%) |  |
| Car | 22 (26%) | 21 (37%) | 1 (3.4%) |  |
| Motorcycle taxi | 1 (1.2%) | 1 (1.8%) | 0 (0%) |  |
| Taxi or similar services | 3 (3.5%) | 3 (5.3%) | 0 (0%) |  |
| **Home-work commute time** |  |  |  | 0.013 |
| < 30 min | 13 (16%) | 12 (23%) | 1 (3.6%) |  |
| 30 min – 1h | 17 (21%) | 14 (26%) | 3 (11%) |  |
| 1h – 1h30 | 20 (25%) | 8 (15%) | 12 (43%) |  |
| 1h30 – 2h | 14 (17%) | 8 (15%) | 6 (21%) |  |
| ≥ 2h | 17 (21%) | 11 (21%) | 6 (21%) |  |
| Missing information | 5 | 4 | 1 |  |
| **Private Health Insurance** |  |  |  | <0.001 |
| Yes | 53 (62%) | 44 (77%) | 9 (31%) |  |
| No | 33 (38%) | 13 (23%) | 20 (69%) |  |
| **Tobacco use** |  |  |  | 0.8 |
| No | 74 (86%) | 50 (88%) | 24 (83%) |  |
| Past use | 8 (9.3%) | 5 (8.8%) | 3 (10%) |  |
| Current use | 4 (4.7%) | 2 (3.5%) | 2 (6.9%) |  |
| **Height (m)** | 1.65 (1.59, 1.70) | 1.65 (1.59, 1.69) | 1.68 (1.60, 1.73) | 0.4 |
| Missing information | 1 | 1 | 0 |  |
| **Weight (kg)** | 72 (66, 85) | 70 (65, 85) | 80 (67, 90) | 0.2 |
| Missing information | 1 | 1 | 0 |  |
| **BMI (kg/m<sup>2</sup>)** | 26.3 (24.0, 30.0) | 26.3 (24.3, 29.4) | 26.8 (23.8, 31.6) | 0.7 |
| Missing information | 1 | 1 | 0 |  |
| **Diabetes** |  |  |  | 0.3 |
| Yes | 13 (15%) | 7 (12%) | 6 (21%) |  |
| No | 73 (85%) | 50 (88%) | 23 (79%) |  |
| **Dyslipidemia** |  |  |  | 0.068 |
| Yes | 29 (34%) | 23 (40%) | 6 (21%) |  |
| No | 57 (66%) | 34 (60%) | 23 (79%) |  |
| **Cancer** |  |  |  | >0.9 |
| Yes | 6 (7.0%) | 4 (7.0%) | 2 (6.9%) |  |
| No | 80 (93%) | 53 (93%) | 27 (93%) |  |
| **COPD** |  |  |  | >0.9 |
| Yes | 3 (3.5%) | 2 (3.5%) | 1 (3.4%) |  |
| No | 83 (97%) | 55 (96%) | 28 (97%) |  |
| **Asma** |  |  |  | 0.4 |
| Yes | 6 (7.0%) | 3 (5.3%) | 3 (10%) |  |
| No | 80 (93%) | 54 (95%) | 26 (90%) |  |
|  | Overall<br>N = 86<sup>1</sup> | Permanent staff<br>N = 57<sup>1</sup> | Contract staff<br>N = 21<sup>1</sup> |  |
| **Rheumatic conditions** |  |  |  | 0.3 |
| Yes | 1 (1.2%) | 0 (0%) | 1 (3.4%) |  |
| No | 85 (99%) | 57 (100%) | 28 (97%) |  |
| **Renal diseases** |  |  |  | >0.9 |
| Yes | 2 (2.3%) | 1 (1.8%) | 1 (3.4%) |  |
| No | 84 (98%) | 56 (98%) | 28 (97%) |  |
| **No pre-existing condition** |  |  |  | 0.14 |
| Yes | 62 (72%) | 44 (77%) | 18 (62%) |  |
| No | 24 (28%) | 13 (23%) | 11 (38%) |  |
| **Multimorbidity (≥2 conditions)** |  |  |  | 0.3 |
| Yes | 5 (5.8%) | 2 (3.5%) | 3 (10%) |  |
| No | 81 (94%) | 55 (96%) | 26 (90%) |  |

<sup>1</sup>Median (Q1, Q3); n (%)
<sup>2</sup>Wilcoxon rank sum test; Fisher's exact test; Pearson's chi-squared test;
Inquired comorbidities with zero answers: rheumatic fever, coronary disease, stroke, and cirrhosis/hepatitis. COPD = chronic obstructive pulmonary disease; BMI = body mass index; R\$ = Brazilian currency (Real). Eight respondents did not answer the professional ties and were not included in the hypothesis tests analysis.

Most of the respondents had private health insurance. In addition, most of them were either overweight or obese according to self-reported weight and height (BMI), on the other hand, the minority reported tobacco use or other morbidities, or multiborbidity. Nevertheless, the prevalence of diabetes was relatively higher, and hypercholesterolemia was relatively lower when compared to the overall population prevalence. (Table 1)

The self-reported health status was usually moderate or good. However, several findings indicate some sort of self-reported symptom or disability. Twenty-five percent of respondents had some mental symptoms, and 21% of the respondents had some disability due to headaches, and burnout scores were nearly all in the second quarter, mostly due to work and personal dimensions. Nearly 10% of respondents had a history of harmful drinking habits or drinking dependence. There was a significant difference in quality of life in the environment domain, where the permanent staff had higher scores. Nevertheless, most respondents reported being happy, with good quality-of-life and spirituality scores, as these were mostly in the upper third range. The perception of institutional support was usually moderate, and there seems to be no difference among staff. (Table 2)

**Table 2.**
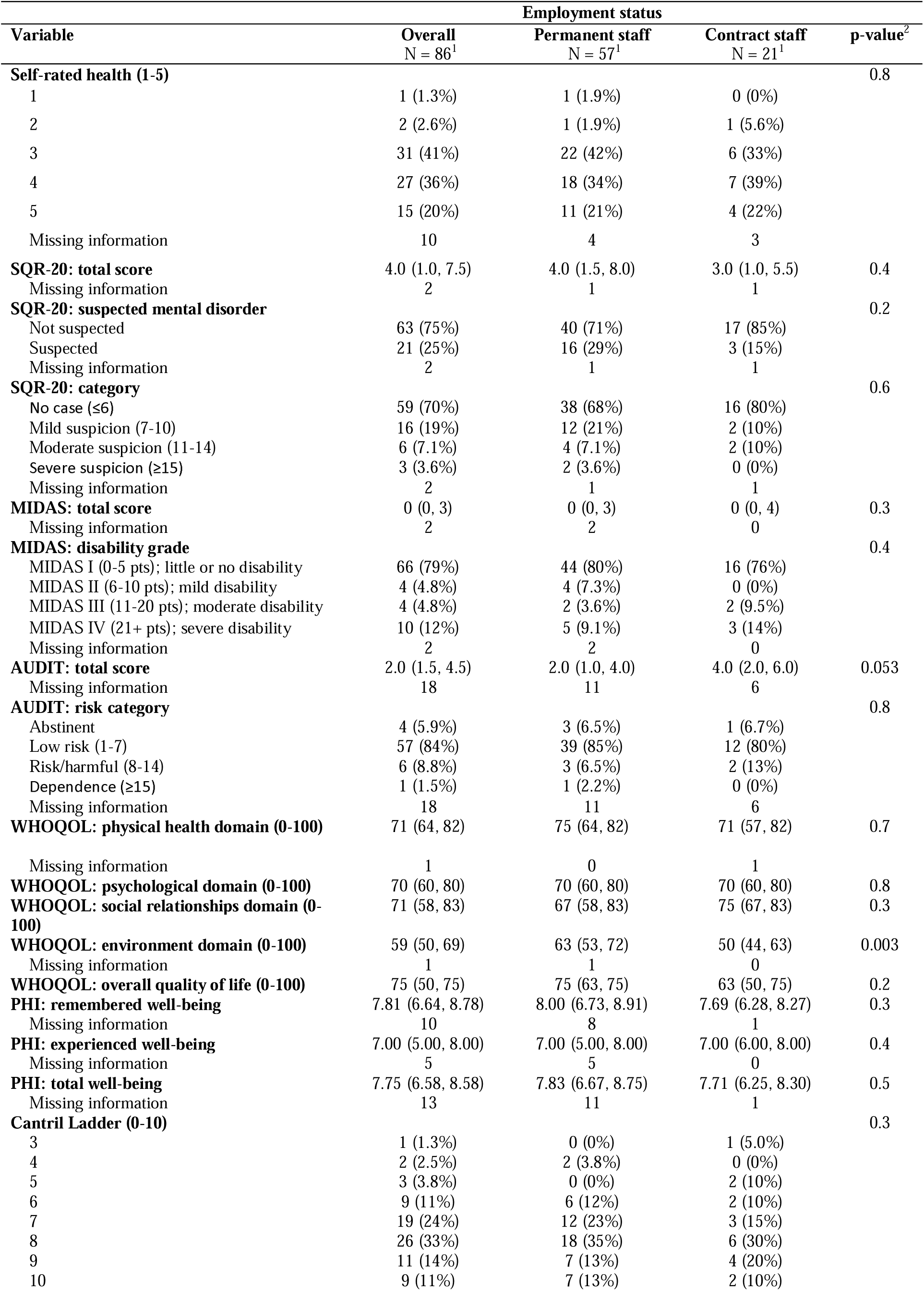
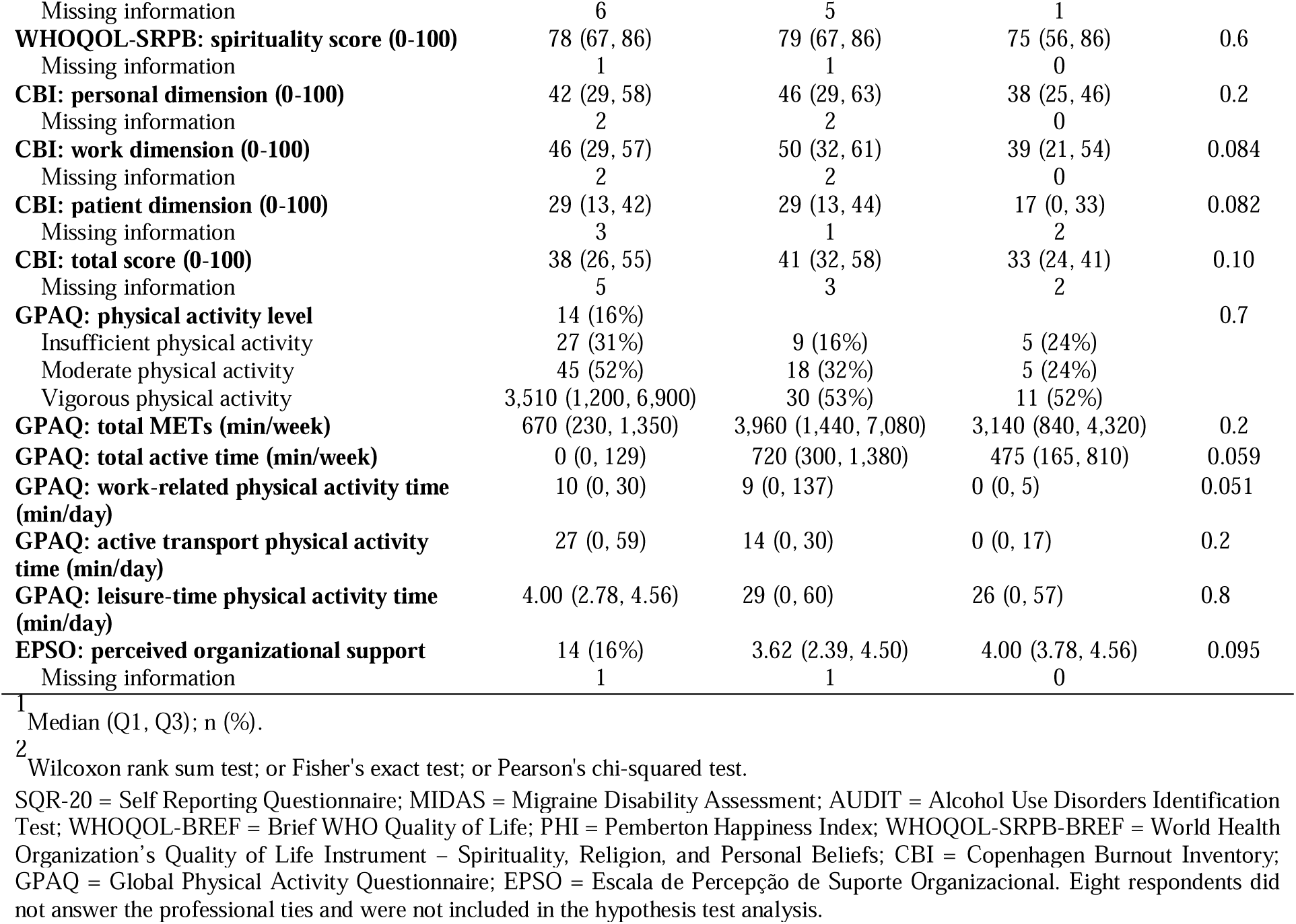
Overall health status measures: total scores from several validated instruments by hospital staff employment relationship.

Nearly half of respondents reported vigorous physical activity, and nearly one-third reported moderate activity, with a median of 670 min/week dedicated to exercise and a median of 3,510 total MET-min/week. Regarding physical activity, 53% of respondents were classified as having vigorous physical activity, 29% as having moderate physical activity, and 18% as having insufficient physical activity. The total median daily time, the work-related physical activity, active transportation, and leisure time spent with physical activity were always higher among the permanent staff, however, there were no statistically significant differences. (Table 2)

The correlation matrix identified a few interesting findings in the overall health assessment. For example, the SRQ-20 score (mental symptoms) was positively correlated with MIDAS (headache) and CBI (burnout) and negatively correlated with quality of life, happiness, spirituality, and perception of institutional support. AUDIT scores (alcoholism) were positively correlated with CBI (burnout) and GPAQ (exercise) in the working dimension; on the other hand, AUDIT was negatively associated with happiness, quality of life, and spirituality. Many dimensions of quality of life were positively correlated with happiness, spirituality, some exercise dimensions, and EPSO (perceptions of institutional support), and negatively correlated with burnout. Happiness was positively correlated with spirituality, exercise in the leisure dimension, and with EPSO scores. At last, CBI (burnout) scores were negatively correlated with EPSO (perceptions of institutional support). (Figure 2)

**Figure 2.**
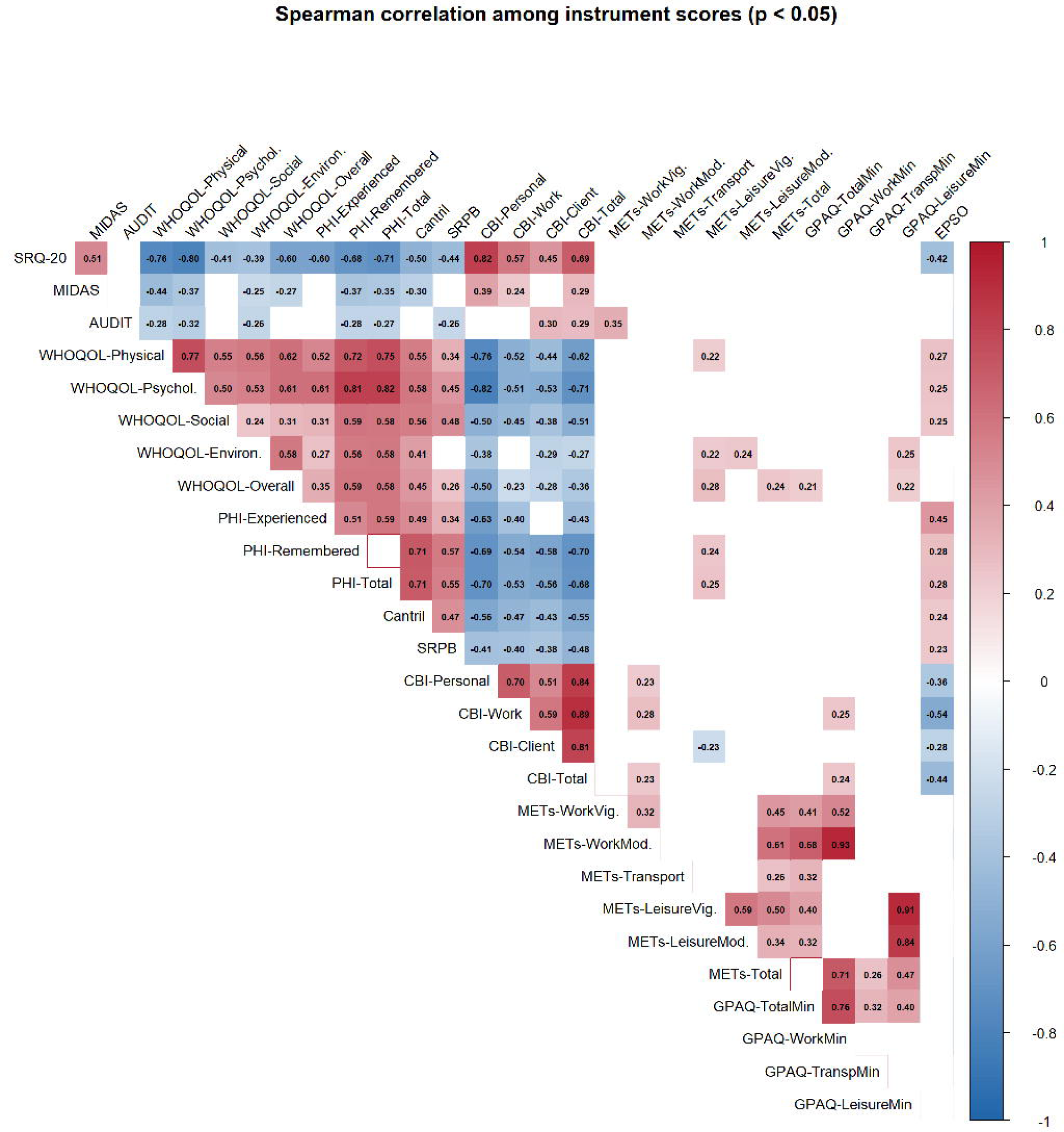
Spearman correlation heatmap of the study health scales. Abbreviations: ICF, Informed Consent Form; SRQ-20, Self-Reporting Questionnaire; MIDAS, Migraine Disability Assessment; AUDIT, Alcohol Use Disorders Identification Test; WHOQOL-BREF, World Health Organization Quality of Life Instrument, abbreviated version; PHI, Pemberton Happiness Index; WHOQOL-SRPB-BREF, WHOQOL Spirituality, Religiousness, and Personal Beliefs module, abbreviated version; CBI, Copenhagen Burnout Inventory; GPAQ, Global Physical Activity Questionnaire; EPSO, Perceived Organizational Support Scale. Correlation coefficients are color-coded from blue (negative) to red (positive), with stronger colors indicating stronger associations. Only significant correlations (<0.05) are shown.

## DISCUSSION

The main results to be discussed are that the project assessed the overall health status of one big metropolitan public hospital; therefore, the set of instruments can be used as a tool to periodically have a situation diagnosis of the global health of the healthcare professionals.

Current strong literature evidence points to physical exercise being most consistently linked to lower stress, better quality of life, and lower depressive burden.^22–25^ There is also evidence that physical activity is positively associated with quality of life among healthcare workers and related populations, particularly in the domains of mental health, vitality, and social functioning.^26–29^ In addition, there is little evidence pointing to exercise improving depression-related and some alcohol-related outcomes; however, evidence for burnout, anxiety, spirituality, and migraine is limited, more mixed, or indirect.^30–33^

Professionals in hospital and administrative environments face specific challenges, such as sedentary work and imbalanced effort-reward ratios, which influence their health outcomes, with evidence of an inverse relationship between exercise and mental symptoms.^34,35^ Physical exercise often intersects with these domains by fostering a sense of purpose, mindfulness, and connection to one’s body and environment. Studies suggest that physical activity can enhance spiritual well-being by providing opportunities for reflection, community engagement, and the experience of “flow” states.^36^ Promoting physical exercise is established as a viable strategy for maintaining work performance and improving health among professionals.^37^ For healthcare workers, teachers, and bankers, physical activity levels are critical factors in mitigating the risk of obesity and sedentary-related health issues.^35^

This relationship is not merely correlational; systematic reviews indicate that exercise interventions lead to measurable improvements in health-related quality of life (HRQoL) by reducing symptom burden and enhancing functional capacity.^38^ In office-based populations, which share similarities with administrative hospital staff, physical exercise has been shown to counteract the negative impacts of prolonged sitting and high workload, thereby preserving HRQoL.^39^

Research with older adults found physical exercise positively correlated with spiritual well-being, an effect partly mediated by self-care ability.^40^ A path-analysis study similarly found that spirituality and physical activity both influence quality of life via self-efficacy, with spirituality showing its strongest effects on mental health status.^41^ A Brazilian study using the WHOQOL-SRPB directly (alongside WHOQOL-Old, anxiety, and depression measures) in 690 older adults found high physical activity levels co-occurred with high spirituality scores (99.6%) and quality of life (73%), with physical activity significantly related to the anxiety/stress/QOL cluster.^42^

Healthcare professionals are disproportionately affected by burnout, anxiety, and depressive symptoms due to high job demands and emotional labor. The Copenhagen Burnout Inventory (CBI) and scales such as the SQR-20, MIDAS, alongside AUDIT and EPSO, are critical for monitoring these risks. Evidence supports an inverse association between physical activity and burnout. Studies have identified physical activity as a key moderator in the relationship between job stress and burnout, with active workers exhibiting lower scores on burnout inventories.^23^ Furthermore, during periods of heightened stress, such as the COVID-19 pandemic, healthcare workers who maintained physical activity levels reported lower prevalence of depressive symptoms and anxiety.^31^

The mechanism involves both physiological adaptations and psychological benefits, including enhanced self-efficacy and mood regulation.^43^ Exercise-based interventions, some as simple as mobile apps, have been shown to improve functioning and reduce depressive symptoms in various clinical and occupational settings, suggesting that promoting physical activity can directly lower scores on distress and burnout scales.^43,44^

There is emerging evidence that physical activity can serve as a constructive coping mechanism, potentially reducing reliance on alcohol for stress relief. While direct causal links in healthcare workers specifically are less extensively documented than in general population studies, broader research indicates that individuals who engage in regular physical activity tend to have lower AUDIT scores.^45^ Meta-analytic evidence supports exercise as an adjunct in alcohol use disorder. One meta-analysis of 17 RCTs (1,905 patients) found exercise significantly reduced AUDIT scores and drinks per day, alongside improvements in VO2max, anxiety, depression, and stress.^32^ A second meta-analysis similarly found exercise reduced drinking volume and improved fitness, though no significant effect on binge drinking specifically.^46^

Aerobic exercise is also associated with reduced migraine burden. Aerobic exercise shows effects comparable to preventive medication for reducing monthly migraine days.^47^ A population-based cohort found that physically active migraine patients had lower MIDAS scores and better life satisfaction, with physical activity independently associated with reduced disability, even after adjustment.^48^

The convergence of evidence suggests a synergistic relationship among these health dimensions. Promoting physical exercise does not operate in isolation; it creates a positive ripple effect across the biopsychosocial spectrum. By improving physical health (WHOQOL-Physical), exercise enhances mental resilience (lowering CBI, SQR-20, and MIDAS scores), reduces maladaptive coping mechanisms like alcohol use (lowering AUDIT scores), and fosters a deeper sense of personal and spiritual well-being (enhancing WHOQOL-SRPB and PHI scores). For hospital administrators and health professionals, implementing structured physical activity programs, such as workplace wellness initiatives or accessible fitness facilities, can yield substantial returns in terms of reduced burnout, improved job satisfaction, and better overall quality of life.^37^

## CONCLUSIONS

Currently, the project staff is convinced that a set of feasible instruments is available to check the hospital staff’s overall health regarding several dimensions. The project’s data collection first wave was considered successful, despite a small sample size and varying degrees of completeness of these instruments. The following waves (originally planned to be once a year) are currently being planned with the same instruments, with data collectors using tablets to conduct interviews, in addition to the web survey. Additional strategies to engage participants were considered and are in the plan: (a) use of social media specifically to project to advertise the data collection and disseminate key results; (b) use of IT support to configure the workstations’ opening screen to advertise the project; (c) open a dashboard to show the community participation engagement of different hospital sectors in real time; (d) repeat some of the considered successful strategies, such as the community walk event.

The effort of implementing the project as a “situation diagnosis” has already fostered an awareness among hospital staff that health promotion can be addressed within the workplace; consequently, there is a plan to build an on-site gym for employees even before the first wave results become fully available and to promote on-site catch-up vaccination campaigns.

## AKNOWLEDGEMENTS

Nothing to declare

## LIST OF ABBREVIATIONS.

*AUDIT*: Alcohol Use Disorders Identification Test
*IBGE*: Instituto Brasileiro de Geografia e Estatística (Brazilian Institute of Geography and Statistics)
*BMI*: Body Mass Index
*CAAE*: Certificado de Apresentação de Apreciação Ética (Certificate of Presentation for Ethical Consideration)
*CBI*: Copenhagen Burnout Inventory
*CNS*: Conselho Nacional de Saúde (Brazilian National Health Council)
*EPSO*: Escala de Percepção de Suporte Organizacional (Organizational Support Perception Scale)
*GPAQ*: Global Physical Activity Questionnaire
*HFSE / HUSE*: Hospital Federal dos Servidores do Estado / Hospital Universitário dos Servidores do Estado
*HRQoL*: Health-Related Quality of Life *MET:* Metabolic Equivalent of Task
*MIDAS*: Migraine Disability Assessment
*PHI*: Pemberton Happiness Index
*QoL*: Quality of Life
*RCT*: Randomized Controlled Trial
*REDCap*: Research Electronic Data Capture
*SRQ-20*: Self-Reporting Questionnaire-20
*WHO*: World Health Organization
*WHOQOL-BREF*: World Health Organization Quality of Life - Abbreviated Version
*WHOQOL-SRPB-BREF*: World Health Organization Quality of Life – Spirituality, Religiousness and Personal Beliefs (Abbreviated)
*VO2max*: Maximal Oxygen Consumption

## DATA AVAILABILITY STATEMENT

The data can be obtained from the corresponding author upon reasonable request.

## AUTHORS’ CONTRIBUTIONS

Rodrigo Teixeira Amancio da Silva - conceptualization, investigation, writing - review & editing Letícia Nascimento Cruz - investigation and coordination. writing - review

Rochelle Dantas - investigation, coordination, writing - review Amanda de Araujo Batista da Silva - data analysis, writing - review Marcia Pereira Gomes - study coordination, writing - review

Pedro Emmanuel Alvarenga Americano do Brasil - conceptualization, methodology, data curation, visualization, roles/writing - original draft, writing - review & editing

## DECLARATION OF GENERATIVE AI AND AI-ASSISTED TECHNOLOGIES IN THE WRITING PROCESS

During the preparation of this work, the author(s) used Grammarly, Quillbot, and Copilot to check grammar and spelling, reduce word count, and improve reading fluency. After using these tools/services, the author(s) reviewed and edited the content as needed and take full responsibility for the content of the publication.

## Data Availability

All data produced in the present study are available upon reasonable request to the authors

